# Causal Effects of Maternal BMI on Pregnancy Outcomes: A Mendelian Randomisation Study Investigating the Mediating Role of Blood Counts

**DOI:** 10.1101/2025.08.21.25334200

**Authors:** Christopher Flatley, Geng Wang, Alesha Hatton, Kym-Mai Nguyen, Liang-Dar Hwang, Nicole M Warrington

## Abstract

**Background:** Pregnancy requires a delicate balance between the maternal immune system and inflammatory responses. Elevated maternal body mass index (BMI) significantly compromises the immune system and increases systemic inflammation. High maternal BMI is associated with adverse pregnancy outcomes, including an increased risk of both pre-eclampsia and preterm birth, which may be mediated through immune-related blood cell changes.

**Methods:** This study used Mendelian randomisation (MR) to investigate the causal relationship between maternal BMI and pregnancy outcomes, including birth weight, placental weight, gestational duration, and pre-eclampsia. We applied two-step MR to assess whether immune-related blood counts, such as neutrophils, lymphocytes, and platelets, mediate these relationships. Single nucleotide polymorphism (SNP) effect estimates for maternal BMI and pregnancy outcomes were sourced from publicly available genome-wide association studies (GWAS), with pregnancy outcomes partitioned into maternal genetic effects to proxy genetic effects on the intrauterine environment.

**Results:** We found that elevated maternal BMI causally increased placental weight (β_IVW_ = 0.164_sd_, P = 2.92 x 10^-7^) and risk of pre-eclampsia (OR_iVW_ 1.75, P = 6.3 x 10^-30^). The effect of maternal BMI on placental weight was larger than its effect on birth weight. Mediation analysis found no evidence of the involvement of immune-related blood counts in these relationships.

**Conclusions:** Maternal BMI has a significant impact on pregnancy outcomes, particularly by increasing placental weight and the risk of pre-eclampsia. These findings highlight BMI-driven placental adaptations as key contributors to pregnancy complications.

## Introduction

While maternal body mass index (BMI) typically increases during pregnancy, a global trend of higher pre-pregnancy BMI among women of childbearing age has emerged [1, 2]. Elevated maternal BMI is associated with adverse pregnancy outcomes. Pre-pregnancy obesity (defined as BMI greater than 25kg/m^2^) significantly increases the risk of pre-eclampsia (Odds Ratio (OR) 3.01, 95% CI: 2.86, 3.17) [2], fetal macrosomia (OR 2.28, 95% CI: 2.15, 2.41) [1], and preterm birth (1.17, 95% CI: 1.13, 1.21) [1].

Pregnancy relies on a delicate equilibrium between the maternal immune system and inflammatory responses, pivotal for its successful progression from ovulation to labour onset. This process begins with local inflammatory reactions triggered by blastocyst-endometrial contact, followed by decidualisation, which starts during the menstrual cycle. Decidualisation allows endometrial stromal cells to evaluate blastocyst quality through changes in their secretory profile [3, 4]. This is followed by the release of pro-implantation factors and immunological regulation, particularly involving lymphocytes and neutrophils, which are essential for a successful pregnancy [5, 6]. Though physiological increases in white blood counts are common during pregnancy, excessive alterations may precipitate pregnancy complications [7–9].

Elevated BMI is associated with systemic chronic inflammation [1, 10], leading to shifts in immune-related blood cell populations, including monocytes, neutrophils, and lymphocytes [11]. Macrophages, derived from monocytes, can constitute up to 40% of obese adipose tissue compared to less than 10% in individuals with a normal BMI [12]. Neutrophils, the most abundant white blood cells (WBCs), are the first responders in the inflammatory response. In obese patients, they infiltrate adipose tissue (unlike in lean individuals, where they are absent from adipose tissue during hemostasis), and therefore elevated neutrophil counts are exhibited, which release significantly more pro-inflammatory mediators [13].

Leukocytosis, a higher-than-normal WBC count, is a normal physiological adaptation during pregnancy. However, increased WBC counts in pregnancy have been associated with pre-eclampsia (OR 1.14, 95% CI: 1.47, 1.64), preterm birth (OR 1.12, 95% CI: 1.06, 1.18), and low birth weight (OR 1.12, 95% CI: 1.08, 1.16) [8]. In individuals with pre-pregnancy obesity, chronic low-grade inflammation may further amplify leukocytosis, potentially exacerbating the risk of these adverse pregnancy outcomes [8, 12].

The pathophysiology of adverse pregnancy outcomes is intricate and multifaceted. Gaining insight into their aetiology and devising strategies to mitigate these outcomes necessitates understanding the causal pathways leading to these outcomes and identifying potentially modifiable risk factors. However, observational association studies are hindered by confounding and reverse causation, which weakens their ability to provide strong evidence of causality. Mendelian randomisation (MR) is a method to infer causal relationships between a modifiable exposure and a relevant disease or trait using genetic variants (typically single nucleotide polymorphisms [SNPs]) as instrumental variables [14, 15]. At the core of MR lies three assumptions that must be met to have valid instrumental variables: the genetic variants must be associated with the modifiable exposure (relevance), there are no confounding variables (measured or unmeasured) between the genetic variants and the outcome (independence), and the genetic variants exclusively impact the outcome through the modifiable exposure (exclusion restriction) [16, 17]. The most commonly used framework, known as two-sample MR, uses SNP-exposure and SNP-outcome estimates from two separate samples (often the largest genome-wide association studies of the traits) to estimate the causal effect of the exposure on the outcome. Two-step MR allows for the investigation of mediating effects of an intermediate variable in a causal modelling framework [18, 19].

Pre-eclampsia has been linked to both cardiometabolic dysregulation and altered systemic inflammatory responses in pregnancy [8, 12, 20, 21]. A recent MR study demonstrated that elevated BMI is causally associated with an increased risk of pre-eclampsia (OR 1.68, 95% CI: 1.46, 1.94, P = 8.74 × 10⁻¹³) [21]. Tyrrell *et al.* (2016) demonstrated a causal association between maternal BMI and offspring birth weight, showing that a one standard deviation increase in maternal BMI was associated with a 55-gram increase in birth weight (95% CI: 17, 93 g) [22]. A MR study investigating BMI and blood traits found a negative causal association between BMI and both WBC and platelet count[23]. Additionally, lymphocyte count is causally associated with pre-eclampsia (OR 1.10, 95% CI: 1.01, 1.21) [24]. Finally, a study examining various leukocyte subsets identified causal relationships between specific immune cell populations and both birth weight and risk of preterm birth [25]. These findings indicate complex relationships between maternal BMI, blood counts and various pregnancy outcomes.

The causal relationships identified in these previous MR studies lend themselves to the hypothesis that blood counts may play a mediatory role in the relationship between maternal BMI and pregnancy outcomes. Therefore, this study aims to use MR to investigate the causal relationship between maternal BMI and pregnancy outcomes, including birth weight, placental weight, gestational duration and pre-eclampsia and hypertensive disorders. If a causal relationship exists, we will use a two-step MR to investigate whether blood counts - including basophil, eosinophil, lymphocyte, monocyte, neutrophil, and platelet counts - mediate the relationship.

## Methods

### Instrumental Variables

#### Body Mass Index

We extracted summary data from the largest European genome-wide association study (GWAS) on BMI to date, which included approximately 700,000 individuals [26]. This meta-analysis combined GWAS results for BMI from the UK Biobank (UKBB) and the GIANT consortium [26]. To identify SNPs to use as instrumental variables, clumping was performed using the TwoSample-MR package (Version 0.6.4) in R (Version 4.2.1) on the GWAS summary statistics, using P < 5 x 10^-8^, r^2^ = 0.001 and within a 10,000kb window. This identified 521 independently associated genetic variants for BMI; however, due to some missing data in the blood count and pregnancy outcome GWAS, the number of genetic variants used as instruments for BMI in each of the analyses, along with the mean F-statistic, can be found in **Supplemental Table 1.**

### Mediator

#### Blood Counts

Summary statistics for SNPs associated with blood count phenotypes (basophil, eosinophil, monocyte, lymphocyte, neutrophil, and platelet counts) were obtained from a large GWAS utilising the UKBB [27]. This study included 408,112 participants of European ancestry. Platelet counts were directly measured in the UKBB as the number of platelets per unit volume of blood using impedance. The remaining blood counts were derived, first as a percentage of WBCs and then converted to absolute counts using the formula [27]:

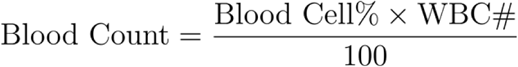

Where WBC# represents the total WBC count, and Blood Cell% represents the percentage of WBCs for that specific blood cell type.

Prior to GWAS analyses, log_10_-transformed blood counts were adjusted for age, age squared, sex, principal components, and cohort-specific covariates, and residuals were inverse-normalised [27].

To perform a two-step MR, two analyses are required using the blood counts. First, SNPs associated with these blood counts were used as instrumental variables to assess relationships between blood counts and pregnancy outcomes. Second, SNP-blood cell count estimates were used to investigate the causal relationship between BMI and blood counts. We employed the same clumping and threshold methods described for the BMI summary statistics to identify independent SNPs to use as instrumental variables for each blood cell count. The number of variants used as instruments for each blood cell count, along with the mean F-statistic, is provided in **Supplemental Table 1**.

### Outcome Variables

Correlated maternal and fetal genomes both influence pregnancy-related measures, and therefore, conditional analyses are required to partition the genetic effect into maternal and fetal-specific components. A method, the weighted linear model (WLM), has been developed that takes the unadjusted genetic effect estimates from GWAS and transforms them into adjusted fetal- and maternal-specific genetic effects [28]. To avoid violating the assumptions underlying MR, the maternal-specific genetic effect was used in the current analyses to proxy the maternal exposures during pregnancy (BMI and blood counts) [29].

#### Birth Weight

Birth weight GWAS summary statistics were obtained for offspring birth weight (n = 270,002) and (own) birth weight (n = 423,683) from Juliusdottir *et al* [30]. First, the DECODE birth weight measures were adjusted for offspring sex, year of birth, gestational age at birth, and maternal age, and then a rank-based inverse normal transformation was applied. A GWAS was then performed using BOLT-LMM (v2.1) [31]. Subsequently, a meta-analysis was performed on both the offspring’s birth weight and own birth weight, combining the DECODE summary statistics with previously published results from the EGG consortium and UKBB participants of European ancestry [30]. To estimate maternal-specific genetic effects on offspring birth weight, we applied a weighted linear model [28] to the summary statistics from this meta-analysis. To achieve this, we merged the offspring birth weight GWAS summary statistics with the fetal (own) birth weight summary statistics and derived the adjusted maternal effect using [28]:

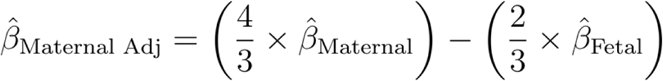

Where β̂_Maternal_ is the SNP effect size from the (unadjusted) GWAS of the maternal genome on offspring birth weight, and β̂_Fetal_ is the SNP effect size from the (unadjusted) GWAS of the fetal genome on their own birth weight. For more information regarding the WLM, see Weighted Linear Model to Partition the SNP Effect Estimates on Birth Weight, in the **Supplementary Methods**.

#### Placental Weight

Placental weight summary statistics were sourced from Beaumont *et al*., who conducted a GWAS on placental weight using fetal (n = 65,405), maternal (n = 61,228), and paternal (n = 52,392) genotypes of European ancestry [32]. The analysis accounted for fetal sex and gestational age, and outcomes were standardised to Z-scores to adjust for variability in collection methods (e.g., trimmed vs. untrimmed placentas). The study included placentas from births between 37 and 43 weeks of gestation, with placental weights ranging from 200 to 1,500 grams. Beaumont *et al*. partitioned their results into maternal-specific effects using the WLM approach described above, which we utilised for this current study [32].

#### Gestational Duration

GWAS of gestational duration was sourced from the fetal genome GWAS by Liu *et al*. [33], (n = 84,689) and the maternal genome GWAS of Solé-Navais *et al.* [34] (n = 195,555). Both GWAS were performed on subjects of European ancestry. The partitioned maternal-specific effect from these GWAS was obtained using the Direct and INdirect effects analysis of Genetic lOci (DINGO) method from Hwang *et al* [35]. This approach partitions the genetic effect into maternal and offspring-specific genetic components in a similar fashion to the WLM approach. Partitioned summary statistics are reported as standardised Z-scores [35].

#### Pre-eclampsia and Hypertensive Disorders of Pregnancy

A pre-eclampsia and hypertensive disorders of pregnancy GWAS of European ancestry was conducted by Tyrmi *et al.* (cases = 15,200, controls = 115,007) [36]. Cases for the pre-eclampsia and hypertensive disorders of pregnancy phenotype were based on International Classification of Diseases codes from ICD-10 (O10, O11, O13, O14, 015, O16), ICD-9 (642) and ICD-8 (63701, 63703, 63704, 63709, 63710, 63799, 66120), and parous women without these codes were classified as controls [36]. We used the SNP-effect sizes from this GWAS for our MR analyses.

### Two-Sample Mendelian Randomisation

Two-sample MR estimates a causal effect of an exposure on an outcome by calculating a Wald ratio for each instrumental variable (here, we use SNPs), which is the ratio of the SNP-outcome association to the SNP-exposure association [37]. These Wald ratios are then meta-analysed using a multiplicative random effects inverse variance weighted (IVW) approach to provide an overall causal estimate [38, 39]. We performed IVW analyses to assess whether there is a causal relationship between maternal BMI, blood counts and pregnancy outcomes (birthweight, placental weight, gestational duration, pre-eclampsia).

Horizontal pleiotropy, where the instrumental variable is related to the outcome through a path other than the exposure, is a concern in MR studies as it violates the exclusion restriction assumption [37]. Therefore, in addition to the IVW method, we also performed sensitivity analyses using multiple pleiotropy-robust MR models (MR-Egger, weighted median, simple mode, weighted mode) that have varying assumptions regarding horizontal pleiotropy[16]. Cochran’s Q test was used to assess the heterogeneity of causal estimates across the different instrument variables.

All two-sample MR models were performed in R (Version 4.2.1) using the TwoSampleMR Package (Version 0.6.4) [40]. We used a threshold of P < 0.05 to determine putative causal relationships to take forward to the two-step MR mediation analysis. A Bonferroni correction was applied to determine causal significance in the two-sample MR analysis of pregnancy outcomes, accounting for the 7 exposures (blood counts and BMI), setting the threshold at P < 0.007.

### Two-Step Mendelian Randomisation

Two-step MR extends the classical two-sample MR framework by incorporating an additional step to assess mediation [19]. After confirming that a causal relationship exists between the exposure (maternal BMI) and outcome (pregnancy outcomes; Figure 1A), two-step MR tests if a causal relationship exists between the exposure of interest (e.g., BMI) and a mediator (e.g. blood counts; Figure 1B), and between the mediator (blood counts) and the outcome of interest (e.g., pregnancy outcomes; Figure 1C).

**Figure 1:**
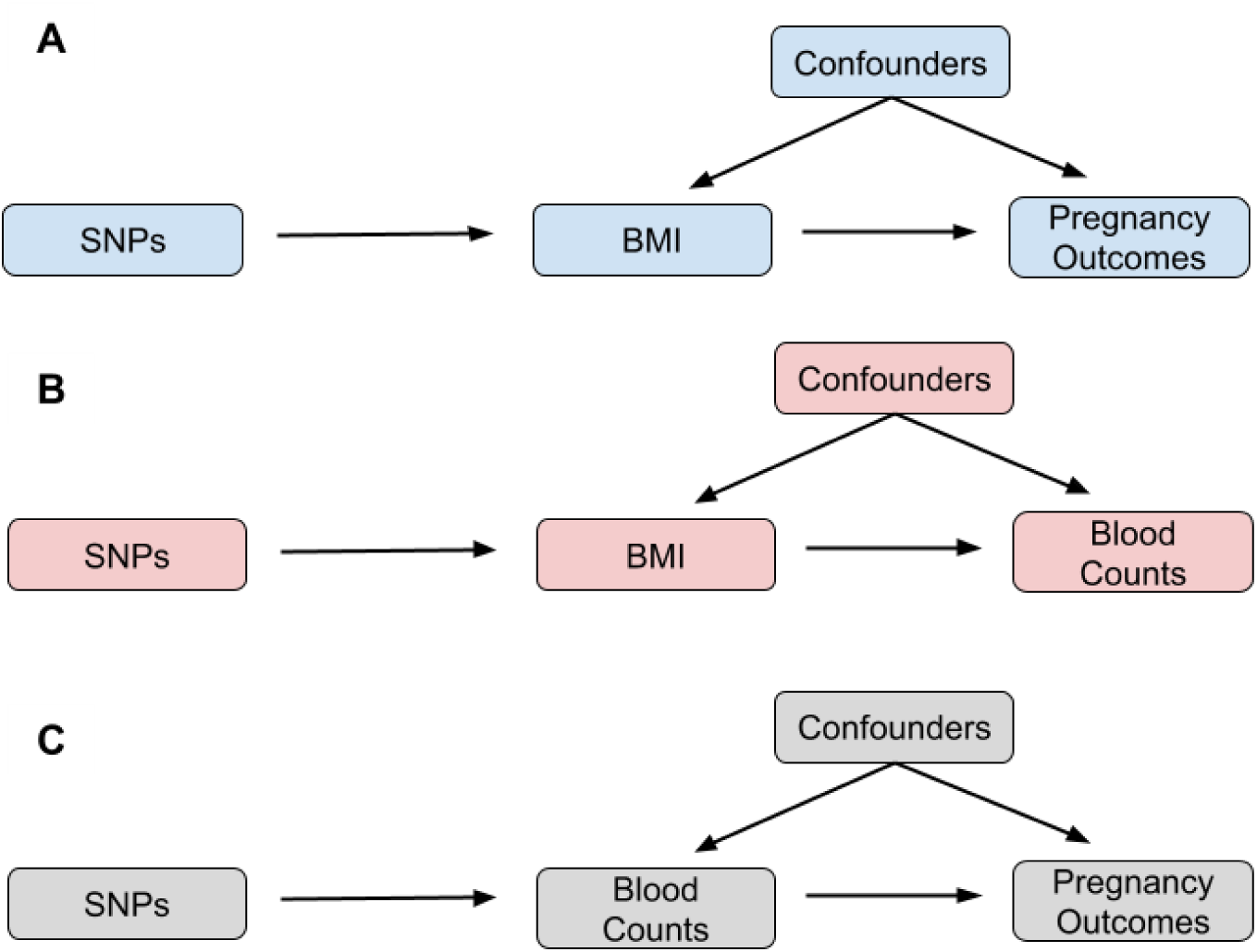
Putative causal effects to be investigated using two-step Mendelian randomisation. SNPs: Single Nucleotide Polymorphisms, BMI: Maternal Body Mass Index

In mediation analysis, three statistics are of interest: the total effect, the direct effect, and the indirect effect. The total effect represents the univariate causal effect of BMI on pregnancy outcomes as derived from the two-sample MR (Figure 1A). The indirect effect captures the influence of BMI on pregnancy outcomes, which occurs solely through the mediating blood count trait. The direct effect is the estimated effect of BMI on pregnancy outcomes after accounting for the indirect effect. To estimate the indirect mediation effect using two-step MR, we apply the product of coefficient method, which multiplies the causal effect of the exposure on the mediator (Figure 1B) by the causal effect of the mediator on the outcome (Figure 1C). To isolate the direct effect of BMI on pregnancy outcomes, we can subtract the indirect effect from the total effect. The standard error for the indirect effect was derived using the delta method[41]:

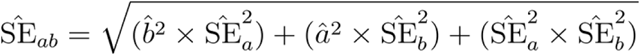

Where the indirect effect:

- *â* is the estimated causal effect of the exposure (BMI) on the mediator (blood count).
- *b̂* is the estimated causal effect of the mediator (blood count) on the outcome (pregnancy outcome).
- *SÊ_a_* is the standard error of the estimated causal effect of the exposure on the mediator.
- *SÊ_b_* is the standard error of the estimated causal effect of the mediator on the outcome.

The standard error for the direct effect is taken from the difference of two estimates method[42]:

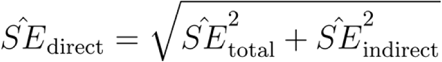

- *SÊ*_total_ is the standard error of the total effect of exposure on the outcome.
- *SÊ*_indirect_ is the standard error of the indirect effect (see above).

The p-value was calculated as a two-tailed probability from a Z-distribution. We calculated a total, direct and indirect effect if we detected a causal relationship (P < 0.05) between the exposure, mediator and outcome (i.e. if we detected a causal effect between BMI and birth weight, BMI and eosinophil counts, and eosinophil counts and birth weight then the total, direct and indirect effects were calculated).

## Results

### Effect of BMI on pregnancy outcomes

Higher maternal BMI was found to causally increase offspring birth weight (β_IVW_ = 0.04_sd_, 95% CI: 0.01, 0.07, P = 0.005), but significant heterogeneity was detected (P_het_ = 2.79 x 10^-68^). The 95% confidence intervals around the causal effect estimates using the pleiotropy-robust methods (weighted mode, weighted median, and MR-Egger) overlapped with those from the IVW but were wider and crossed the null (Figure 2, **Supplementary Table 2**).

**Figure 2:**
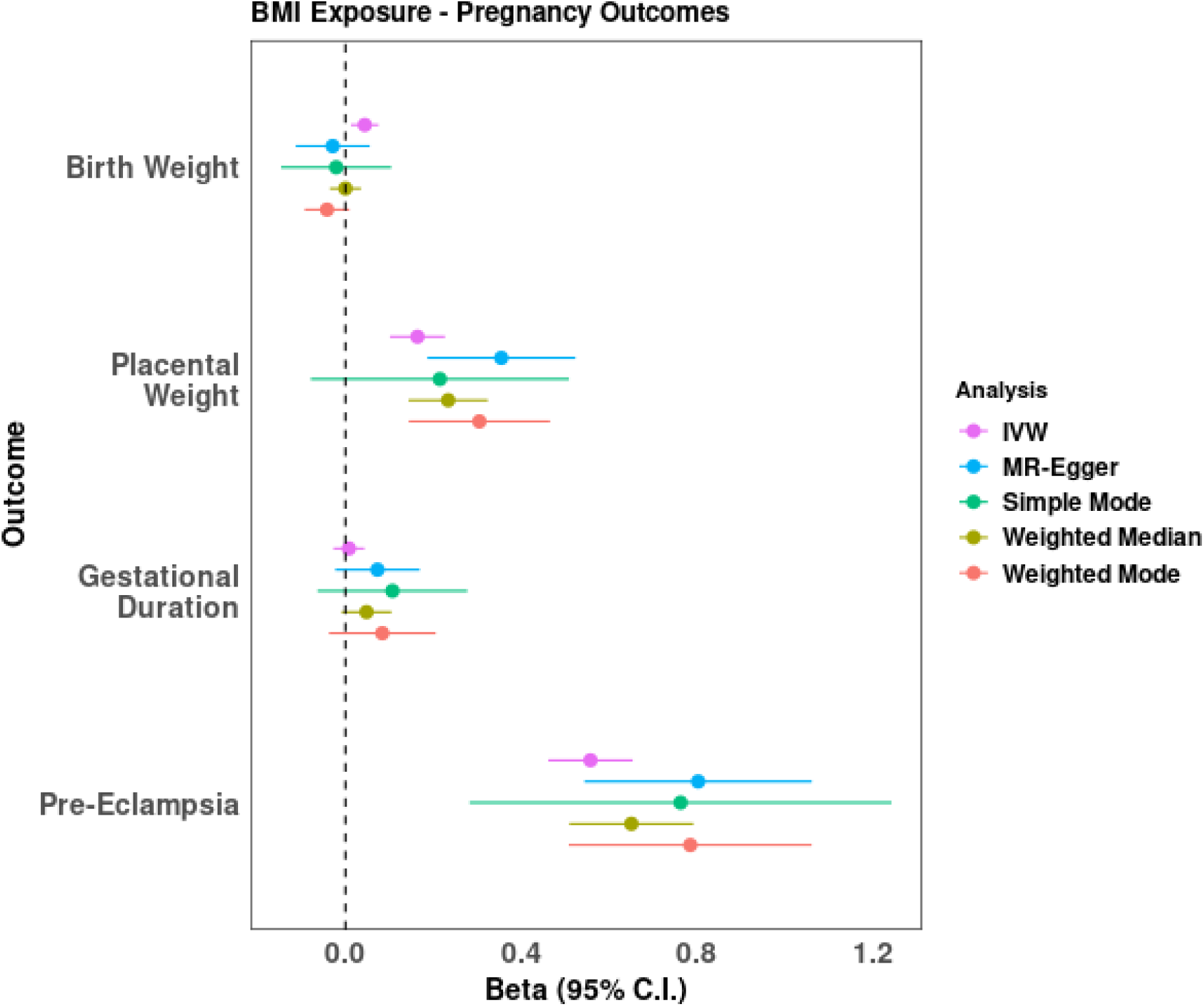
Results from two-sample Mendelian Randomisation analyses of BMI on pregnancy outcomes

IVW analysis indicated a causal association between elevated maternal BMI and increased placental weight (β_IVW_ = 0.16_sd_, 95% CI: 0.10, 0.23, P = 2.92 x 10^-7^). Similar to birth weight, significant heterogeneity was detected (P_he t_= 7.47 x 10^-7^), and the MR Egger intercept indicated directional pleiotropy (P = 0.02; Supplementary Table 2). The 95% confidence intervals from the pleiotropy-robust methods overlapped the 95% confidence intervals from the IVW analyses (Figure 2, **Supplementary Table 2**).

No significant causal relationship was observed between maternal BMI and gestational duration (β_IVW_ = 0.01_sd_, 95% CI: -0.03, 0.04, P = 0.67). Again, significant heterogeneity was detected (P_het_ = 0.03), and the pleiotropy-robust methods also show no evidence of a causal effect (Figure 2, **Supplementary Table 2**).

Finally, a causal association was observed between maternal BMI and risk of pre-eclampsia and hypertensive disorders of pregnancy (OR 1.75, 95% CI: 1.59, 1.92, P = 6.33 x 10^-30^), indicating that a higher BMI increased the risk of developing pre-eclampsia and hypertensive disorders while pregnant. While there was significant heterogeneity (P_het_ = 1.18 x 10^-6^). The MR-Egger intercept suggests directional pleiotropy (P = 0.05), and the pleiotropy-robust methods support this finding of causality (Figure 2, **Supplementary Table 2**).

### Effect of BMI on blood counts

The IVW results provided evidence that higher BMI causes a lower basophil count (β_IVW_ = -0.02_sd_, 95% CI: -0.05, -0.002, P = 0.03), eosinophil count (β_IVW_ = -0.04_sd_, 95% CI: -0.08, -0.004, P = 0.03), monocyte count (β_IVW_ = -0.07_sd_, 95% CI: -0.10, -0.03, P = 9.80 x 10^-5^) and platelet counts (β_IVW_ = -0.07_sd_, 95% CI: -0.12, -0.03, P = 0.001). We detected significant heterogeneity for all of these causal effects, and the estimated causal effects attenuated towards the null when using approaches more robust to pleiotropy (**Supplementary Figure 1, Supplementary Table 3**). BMI showed no causal relationship with lymphocyte count (P = 0.23) or neutrophil counts (P = 0.30, **Supplementary Figure 1, Supplementary Table 3**).

### Effect of Blood Counts on Birth Weight

Of the blood counts, both eosinophil count (β_IVW_ = -0.03_sd_, 95% CI: -0.05, -0.01, P = 0.01) and lymphocyte count (β_IVW_ = -0.04_sd_, 95% CI: -0.06, -0.02, P = 0.001) were found to have a causal relationship with lower birth weight. Significant heterogeneity was again detected for both causal relationships (P_het_ = 5.62 x 10^-30^, P_het_ = 4.47 x 10^-35^ for eosinophil and lymphocyte counts, respectively). The pleiotropic-robust methods showed consistent results with slightly larger standard errors (**Supplementary Figure 1, Supplementary Table 4**). No other blood counts were found to have causal relationships with birth weight. (**Supplementary Figure 1, Supplementary Table 4**).

### Effect of Blood Counts on Placental Weight

No causal relationships were observed between any of the blood counts and placental weight (**Supplementary Figure 1, Supplementary Table 5**).

### Effect of Blood Counts on Gestational Duration

IVW analysis suggested a causal association between eosinophil count and gestational duration (β_IVW_ = -0.04_sd_, 95% CI: -0.07, -0.01, P = 0.006, P_het_ = 0.014), with the 95% confidence intervals around the causal effect estimates using the pleiotropy-robust methods overlapping with those from the IVW. No significant causal relationship was observed between any other blood count and gestational duration (**Supplementary Figure 1, Supplementary Table 6**).

### Effect of Blood Counts on Pre-eclampsia and hypertensive disorders of pregnancy

Higher neutrophil counts were found to have a protective causal effect on the risk of pre-eclampsia and hypertensive disorders (OR 0.89, 95% CI: 0.82, 0.98, P = 0.012). Heterogeneity was again detected (P_het_ = 9.02 x 10^-7^). However, the 95% confidence intervals around the causal effect estimates using the pleiotropy-robust methods overlapped with those from the IVW but were wider. No other blood counts were found to have a causal relationship with pre-eclampsia and hypertensive disorders (**Supplementary Figure 1, Supplementary Table 7**).

### Two-Step Mendelian Randomisation and Mediation Analysis

In the two-step MR mediation analysis, the only combination that indicated a causal relationship between the exposure (BMI), the mediator (blood counts), and one of the pregnancy outcomes in the two-sample MR was between BMI, eosinophil count and birth weight. Therefore, a formal two-step MR was performed only for this combination of traits.

Using two-step MR, the total effect of BMI on birth weight was β = 0.04_sd_ (95% CI: 0.01, 0.07, P = 0.01). There was no evidence of an indirect effect (β = 0.001_sd_, 95% CI: -0.0002, 0.002, P = 0.11). Subsequently, the direct effect of BMI on birth weight (β = 0.04_sd_, 95% CI: 0.01, 0.07, P = 0.01) was very similar to the total effect. (Figure 3)

**Figure 3:**
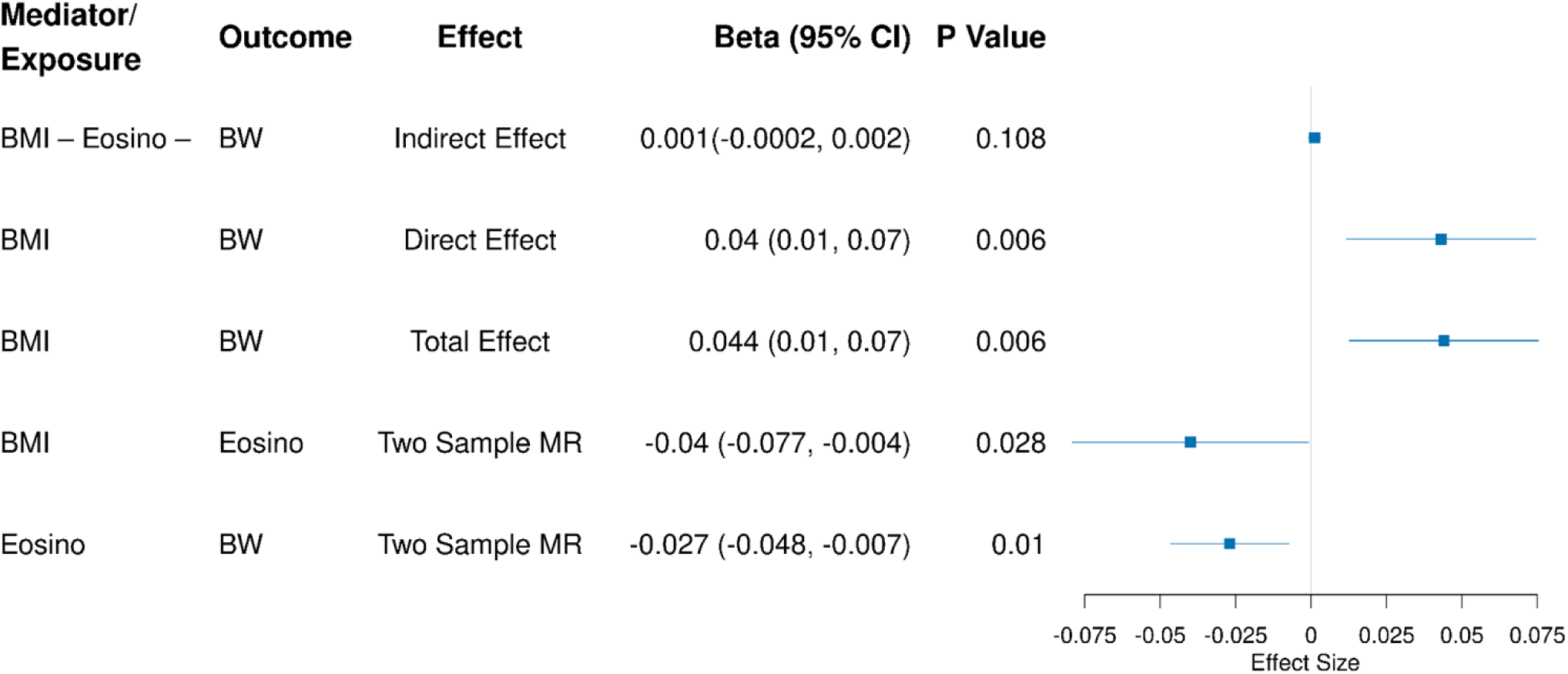
Mediation Effects of Eosinophil Counts on the BMI - Birth Weight Causal Pathway. Eosino: Eosinophil Count; BMI: Maternal Body Mass Index; BW: Birth Weight

## Discussion

We utilised genetic variants associated with BMI to investigate its relationship with pregnancy outcomes and to determine whether this relationship is mediated by blood counts. We observed a causal relationship between maternal BMI and offspring birth weight, similar to but slightly lower than the findings of Tyrrell *et al* [22] (if one standard sd in birth weight is equivalent to 492g [30], then we estimate a 21.6g [95% CI: 6.4, 36.4] increase in birth weight for each kg/m^2^ increase in maternal BMI, whereas Tyrrell *et al.* estimate a 55g [95% CI: 17, 93] increase in birth weight). However, our sensitivity analyses indicate that no causal relationship exists between maternal BMI and birth weight after adjusting for pleiotropy. This could not be tested by Tyrrell and colleagues [22] as they used a genetic score to test their causal relationship, which is not amenable to the pleiotropy robust methods used in the current study. Notably, however, we identified a much stronger causal effect of maternal BMI on placental weight - an association that, to our knowledge, had not been previously assessed using an MR approach. Given the established genetic relationships between birth weight, placental weight, and gestational duration, it is striking that we found no evidence for a causal relationship between maternal BMI and gestational duration. However, we successfully replicated the findings of Ardissino *et al* [21], confirming a causal relationship between maternal BMI and pre-eclampsia.

While observational studies have shown that maternal BMI is associated with both placental weight and birth weight independently [43] and that overweight and obese mothers tend to have disproportionately large placentas relative to birth weight [44], our findings suggest a causal relationship more than five times stronger than its impact on birth weight. Placental function is shaped by maternal metabolic signals, many of which are altered by obesity. Emerging evidence suggests that obesity-driven placental changes mediate its adverse effects on fetal development [45–47]. Maternal obesity-induced epigenetic modifications in the placenta, including altered DNA methylation and hydroxymethylation, likely drive placental overgrowth while impairing nutrient transfer efficiency, explaining why maternal obesity has a greater effect on placental weight than birth weight [48, 49].

Observational studies suggest that elevated BMI induces a pro-inflammatory state characterised by increased leukocyte levels. However, our MR analysis provides causal evidence that higher BMI lowers several of the leukocyte and platelet counts. This replicates previously reported causal associations between maternal BMI and blood count traits [23] and indicates that alternative inflammatory mechanisms may be driving these effects.

Both eosinophil count and lymphocyte count were found to be negatively causally associated with birth weight, with eosinophil count also being negatively causal for gestational duration. The maternal eosinophil count typically decreases slightly or remains stable throughout an uncomplicated pregnancy [50–52]. Nonetheless, several cytokines, particularly interleukin-5, can stimulate eosinophil production, leading to eosinophilia [53, 54]. However, most cases of eosinophilia are hereditary, resulting from an autosomal dominant disorder located in the chromosomal region 5q31-q33 [55]. Elevated eosinophil counts are also associated with a severe form of asthma known as eosinophilic asthma, characterised by the TH2-high asthma endotype [56]. Maternal asthma is the most prevalent chronic condition during pregnancy and has been linked to reduced birth weight in offspring [57, 58].

Lymphocyte counts are generally reported to decrease throughout pregnancy [51, 52]. Lymphocytes, specifically T cells and B cells, are crucial in balancing the maternal immune system during pregnancy to support fetal tolerance [59]. Maternal lymphocytes, in particular decidual natural killer (dNK) cells, are pivotal in the formation of the decidua and the success of implantation [60, 61]. While dNK cells are less cytotoxic, other subsets such as peripheral and endometrial natural killer cells tend to be more cytotoxic, as such, elevated endometrial NK cells have been associated with recurrent pregnancy loss and recurrent implantation failure [62, 63]. This suggests that an imbalance in the maternal immune system lymphocytes negatively affects the maternal revascularisation of maternal spiral arteries and fetal trophoblast invasion of the endometrium [62, 63], resulting in fetal growth restriction or lower birth weight. While elevated lymphocyte counts during pregnancy often result from infection or chronic inflammation, there is also a substantial genetic contribution to an individual’s lymphocyte count [64].

This study found that higher neutrophil counts were associated with a lower risk of pre-eclampsia and hypertensive disorders of pregnancy. While this may seem counterintuitive given the inflammatory nature of pre-eclampsia, evidence suggests that neutrophils play a regulatory role in immune balance rather than solely driving inflammation [6, 65]. A more robust immune response may help mitigate excessive inflammation in pregnancy, potentially explaining our findings. A MR study by Zeng *et al*. (2024) investigating immune dysregulation and inflammatory biomarkers in pre-eclampsia found no association between genetically predicted neutrophil count and pre-eclampsia [24]. However, our study had a larger sample size and a broader phenotype definition, including pre-eclampsia and hypertensive disorders of pregnancy, which increased sensitivity and may have improved our ability to detect an association.

Two-step MR analyses found no evidence that maternal blood counts mediate the effect of maternal BMI on offspring birth weight. This suggests that any inflammatory response linking maternal BMI to birth weight, placental weight, and pre-eclampsia operates independently of the blood count - related inflammatory pathways - which themselves are causally associated with birth weight, gestational duration, and pre-eclampsia. These results underscore a complex interplay: although maternal BMI is causally associated with changes in several blood counts, these alterations do not explain its overall effect on offspring birth weight, placental weight, or pre-eclampsia, indicating that other inflammatory pathways are likely involved.

The strengths of our study lie in the access to large GWAS for all our traits and the partitioned maternal and fetal genetic effects of our pregnancy outcomes. Furthermore, we used several sensitivity analyses with differing assumptions regarding pleiotropy, which gives confidence to our findings. Conversely, our study also has limitations. Horizontal pleiotropy in Mendelian randomisation can only partially be accounted for by methods such as median and mode MR and MR-Egger. The more complex a disease and the more variants used to investigate the causal relationship, the more likely the SNP-outcome relationship is affected by unknown horizontal pleiotropic pathways [66]. Secondly, the lack of population diversity in our chosen GWAS may make our findings non-generalisable to a population outside the European cohorts. Thus, further research is required in other ancestries before conclusions can be made regarding the complex relationship between BMI, immune cell counts, and pregnancy outcomes in other ancestries. Finally, we chose to use a liberal threshold of P < 0.05 for identifying causal relationships to go forward for testing with two-step MR. Therefore, our mediation analysis with eosinophil count and birth weight needs to be replicated.

## Conclusions

This study has shown that elevated maternal BMI has a causal influence on placental weight and that the previously identified relationship with birth weight is likely to be driven by pleiotropy. Additionally, these causal relationships are not mediated via maternal blood counts, indicating that intervening on blood counts during pregnancy is unlikely to influence fetal or placental growth. Continuing to explore the genetic factors, immune cell function, and environmental influences on pregnancy outcomes will be crucial in advancing our knowledge and enhancing maternal and child health.

## Declarations

### Ethics approval and consent to participate

All data used within this study was at the summary level only. As no individual-level data was accessed, no ethical approval was necessary.

### Consent for publication

All authors have agreed to submit and publish this manuscript.

### Availability of data and material

All data used in this study were obtained from publicly available sources. Detailed information on data access and usage is provided in the Methods and Supplementary Materials.

### Author Contributions

C.F: Conception, study design, analysis, interpretation, drafting of the manuscript, and final approval.

G.W: Study design, interpretation, drafting of the manuscript, critical revision, and final approval.

A.H: Study design, interpretation, drafting of the manuscript, critical revision, and final approval.

K.N: Interpretation, drafting of the manuscript, critical revision, and final approval.

L.H: Study design, interpretation, drafting of the manuscript, critical revision, and final approval.

N.W: Conception, study design, interpretation, drafting of the manuscript, critical revision, and final approval.

## Acknowledgments

We gratefully acknowledge the participants and investigators of the genome-wide association studies that made their summary statistics publicly available. Without their contributions, this research would not have been possible.

